# Polygenic Scores for Longitudinal Prediction of Incident Type 2 Diabetes in an Ancestrally and Medically Diverse Primary Care Network

**DOI:** 10.1101/2023.09.08.23295276

**Authors:** Ravi Mandla, Philip Schroeder, Bianca Porneala, Jose C. Florez, James B. Meigs, Josep M. Mercader, Aaron Leong

## Abstract

**OBJECTIVE:** The clinical utility of genetic information for type 2 diabetes (T2D) prediction with polygenic score (PGS) in ancestrally diverse, real-world US healthcare systems is unclear, especially for those at low clinical phenotypic risk for T2D.

**RESEARCH DESIGN AND METHODS:** We tested the association of PGS with T2D incidence in patients followed within a primary care practice network over 16 years in four hypothetical scenarios that varied by clinical data availability (N = 14,712): 1) age and sex, 2) age, sex, BMI, systolic blood pressure, and family history of diabetes; 3) all variables in (2) and random glucose; 4) all variables in (3), HDL, total cholesterol, and triglycerides, combined in a clinical risk score (CRS). To determine whether genetic effects differed by baseline clinical risk, we tested for interaction with the CRS.

**RESULTS:** PGS was associated with incident diabetes in all models. Adjusting for age and sex only, the Hazard Ratio (HR) per PGS standard deviation (SD) was 1.76 (95% CI 1.68, 1.84) and the HR of top 5% of PGS vs interquartile range (IQR) was 2.80 (2.39, 3.28). Adjusting for the CRS, the HR per SD was 1.48 (1.40, 1.57) and HR of top 5% of PGS vs IQR was 2.09 (1.72, 2.55). Genetic effects differed by baseline clinical risk [(PGS-CRS interaction *p*=0.05; CRS below the median: HR 1.60 (1.43, 1.79); CRS above the median: HR 1.45 (1.35, 1.55)].

**CONCLUSIONS:** Genetic information can help identify high-risk patients even among those perceived to be low risk in a clinical evaluation.

## INTRODUCTION

More than a fifth of outpatient visits are for patients with diabetes, largely type two diabetes (T2D), placing a heavy burden on US healthcare systems (1,2). Identifying individuals with elevated risk for T2D is crucial to channel resources towards those most likely to benefit from interventions that prevent or delay diabetes and its complications (3). To assess clinical risk factors for diabetes (e.g., family history for diabetes, obesity, hypertension, hyperlipidemia), clinicians perform a clinical evaluation and order fasting lab tests (e.g., fasting glucose and lipid panel) (4,5). However, patients who do not engage frequently with the healthcare system or are perceived to be low risk at a routine health maintenance exam may not have lab testing or regular follow up visits, limiting accurate risk estimation.

Genetic information, if available, could improve T2D prediction among patients lacking measured clinical risk factors (6). Genome-wide association studies (GWAS) have identified hundreds of unique loci associated with T2D (7), the results of which can be used to calculate polygenic scores (PGS) that model genetic risk independently of established clinical risk factors including family history (8,9). Previous work has evaluated how PGS can be used within healthcare systems (6,10), but analyses have been largely cross-sectional in biobanks of mostly European ancestry, limiting the generalizability of results to a more ancestrally and medically diverse US healthcare system. While PGS have been shown to only modestly improve prediction over traditional clinical risk factors (11,12), the long-term prediction of T2D by PGS in clinical scenarios that vary in clinical information availability in medical records has not been studied.

In a large academic primary care physician (PCP) network affiliated with Mass General Hospital (MGH)(13), 14,712 patients have been genotyped in the Mass General Brigham (MGB) Biobank and followed in the network for up to 16 years. We hypothesized that PGS constructed from their genetic data would be associated with incident diabetes even after adjusting for clinical risk factors available in their electronic health records (EHR). We further hypothesize that PGS would have the most added predictive value in patients with sparse baseline clinical data (i.e., risk factors such as random glucose have not been measured or captured in the EHR).

We constructed four nested cohorts based on hypothetical scenarios by which a patients with genetic data might interact with the healthcare system: (1) a person registered themselves as a patient and provided demographic data; (2) a patient had a visit with a healthcare provider during which their weight, height, and blood pressure were measured and a medical history was taken; (3) a patient had a random glucose measured from a basic lab panel in addition to having a visit with a healthcare provider; (4) a lipid panel, which usually requires the patient to fast overnight, was also performed enabling calculation of the Framingham T2D clinical risk score (CRS) (14). We generated Cox models adjusted for baseline clinical variables in each of these scenarios, and evaluated improvements to T2D incidence prediction when including PGS. We then performed stratified analyses by age, BMI, random glucose, and CRS to evaluate the additional prediction information of PGS for patients that would be considered “lower risk” or “higher risk” at baseline. Finally, as there are few studies on genetic prediction of diabetes-related complications in primary care, we analyzed how PGS can improve prediction of the onset of Coronary Artery Disease (CAD) and Chronic Kidney Disease (CKD), the leading causes of death in people with diabetes (15,16).

## Research Design and Methods

### Data Source and Study Sample

Patients were eligible if they received primary care within the network and had at least two encounters between January 1, 2000, and December 31, 2020. MGB institutional review board approved the study.

### Clinical characteristics

Clinical data was obtained from an EHR repository including MGH outpatient, emergency department, and inpatient visits. Sociodemographic variables included baseline age, self-reported race/ethnicity, educational attainment, and gender. Baseline clinical diagnoses for coronary heart failure, peripheral vascular disease, proteinuria, and hypertension (HTN) were defined with ICD-10 code-based algorithms between January 1, 2000, and up to 6 months after the initial encounter. T2D, CAD, and CKD duration at baseline was calculated from diagnosis date to the date of initial encounter. Mean lab values were extracted between 18 months before the initial encounter and 6 months after the initial encounter, and BMI was extracted anytime in the study.

### T2D Clinical Risk Score

We implemented the Framingham T2D CRS, comprising age, sex, parental history of diabetes, BMI, systolic blood pressure, HDL, total cholesterol, triglyceride, fasting glucose, and waist circumference(14). We used family history as a proxy for parental history. As waist circumference and fasting status for glucose were not available, we removed both variables from the CRS and included random glucose as an independent variable.

### Outcomes

Incidence of T2D, CAD, and CKD were defined with ICD-10 code-based algorithms (Supplemental Table S1) and censored at disease occurrence or the last clinical encounter before December 15, 2021.

### Genotyping Data Preparation

We accessed Multi-Ethnic Genotyping Array (MEGA) genotyping data for 36K participants and Infimum Global Screening Array (GSA) genotyping data for 18K participants from the MGB Biobank then processed by batch. Briefly, the quality control steps included filtering out variants based on MAF levels (<0.5%), missingness (*>*0.05), genotyping batch bias (*P*<5×10^-5^), and Hardy-Weinberg equilibrium (*P*<1×10^-10^) and palindromic single nucleotide variants (AT or CG). Individuals were also removed if their self-described sex did not match their genetic sex or if they had a high ratio of heterozygote variants. Filtering variants based on Hardy-Weinberg equilibrium and participants based on heterozygosity was performed in self-identified race subgroups. These clean datasets were phased using Shapeit4, imputed using the TOPMed r2 reference panel, then union merged together.

To calculate Principal Components (PCs) and genetic ancestry probabilities for individuals in MGBB, we first created an intersection of common (MAF>.5%, genotyping rate >0.95%), independent (R2<0.1) variants from both the HGDP/1000G dataset and MGBB. Next, PCs were calculated among the HGDP/1000G dataset and MGBB was projected into this PC space. Genetic ancestry probabilities were calculated using a random forest logistic model, trained on PCs and continental ancestry data from HGDP/1000G and applied to MGBB. In our analysis, we called an individual as having European ancestry if their European genetic ancestry probability was >0.5.

### Calculation of PGS

We selected large GWAS meta-analyses for our traits of interest of which full summary statistics were available for the calculation of PGS. For T2D we meta-analyzed the published MVP/DIAMANTE meta-analyses results with T2D GWAS from the FINNGEN Biobank r6(17,18).

For CAD we chose the meta-analysis from Nelson et al. of UK Biobank SOFT CAD GWAS with CARDIoGRAMplusC4D 1000 Genomes-based GWAS and the Myocardial Infarction Genetics and CARDIoGRAM (19). For CKD, we downloaded summary statistics from the CKD Gen Consortium(20).

Genome-wide PGS were calculated using PRScs with the provided EUR 1000G HapMap3 LD reference files(21). Posterior weights from PRScs were used to calculate the PGS in the MGB biobank with the PLINK –score function. To account for PGS variability in our multi-ancestry cohort, we implemented a modified PGS adjustment strategy based on previously published methods(10,22). Briefly, we fitted a linear model of each disease-specific PGS against genetic ancestry probabilities. Adjusted PGS were calculated as the residual between the predicted and actual PGS in the entire dataset.

### Primary Analyses

We tested the association of PGS with diabetes incidence in Cox models adjusted for the available clinical variables in each scenario. We corrected for 10 PCs in all models to account for population stratification. As a sensitivity analysis, we considered genetic risk as a categorical measurement whether an individual is within the top 5% of the PGS distribution compared to the PGS interquartile range (IQR) in their scenario. Kaplan-Meier curves and Cox models were generated using the lifelines package in Python(23). Absolute risk was calculated as the predicted probability of having diabetes based on logistic models predicting diabetes status based on PGS and clinical risk factors available in each scenario. Tertiles of PGS were defined among participants within each scenario.

We calculated the change in model performance using the c-index(24) upon adding PGS to several clinical base models: scenario 1) age and sex, scenario 2) age, sex, BMI and systolic BP, scenario 3) age, sex, BMI, systolic blood pressure and random glucose, scenario 4) age, sex, BMI, systolic blood pressure, HDL, total cholesterol, and triglycerides combined into a clinical risk score (CRS) and random glucose. We evaluated the performance of the full CRS in our models using the pre-computed, published weights for each individual variable from the Framingham T2D CRS(14). In a sensitivity analysis, we created separate models built on individual variables without using pre-computed weights in our complete cases. In another sensitivity analysis, we imputed missing information using multivariate feature imputation, imputing missing values using predictions modelled on all available clinical risk factors (Supplemental Table S2).

### Stratified Analyses

For each scenario, we stratified our analyses by the clinical variable available in each scenario that was most associated with diabetes: for scenario 1, we stratified by the recommended age cutoff from the ADA and CDC to commence screening for diabetes at routine health examinations (25,26), 40 years; for scenario 2, the median BMI of our dataset (27.5 kg/m^2^); for scenario 3, a random glucose cutoff of 100 mg/dL (threshold for impaired fasting glucose, presuming blood tests were drawn fasting); for scenario 4, median CRS value.

### Prediction of CKD and CAD as diabetes-related complications

For CKD we used the SCORED CRS, which requires age, sex, and diagnoses of anemia, HTN, diabetes, CHD, congestive heart failure, peripheral vascular disease, and proteinuria diagnoses(27).

For CAD we used the Framingham CAD CRS, which requires age, sex, smoking, total cholesterol, HDL measurements, systolic blood pressure, and whether a patient is undergoing blood pressure treatment(28). We used HTN diagnosis as a substitute for blood pressure treatment.

Similar analyses to those described above for T2D was performed with CAD or CKD as the outcomes in patients with and without T2D. We considered two scenarios for these analyses: (1) a clinical visit without labs and (2) a clinical visit with labs. For CAD, the clinical variables per model per scenario were (1) age, sex, smoking status, and systolic BP and (2) age, sex, smoking status, systolic BP, HDL, and total cholesterol combined into a CRS. For CKD, the clinical variables per model (1) age, sex, diagnosis history, systolic BP, diastolic BP, weight, and HTN, and (2) the risk factors from (1) combined into a CRS.

## RESULTS

We compared the demographic characteristics of genotyped patients in the PCP network (N=15,355), non-genotyped patients (N=269,247) in the network, and genotyped patients in the MGB biobank but not in the network (N=38,107; Supplementary Table S2). Patients with genetic data within the PCP network had a higher proportion of T2D, higher proportion of self-reported non-Hispanic white individuals, and higher educational attainment compared to patients without genetic data and to patients with genetic data but not in the network.

Patients included in Scenarios 3 and 4 with a greater number of clinical risk factors available for analysis at baseline were on average older, had a higher proportion of comorbidities, and a smaller proportion of current smokers compared with patients included in Scenario 1 and Scenario 2. Other baseline characteristics were similar across scenarios (Table 1).

**Table 1:** Patient characteristics in the Massachusetts General Hospital Patient Care Physician Network with genetic data by scenario. Follow-up time was calculated as the length of time between the first diagnosis of diabetes and either January 1, 2000 or a patient’s enrollment into the PCP. Data was left-censored to remove participants with a diagnosis of DM before their start date. Only complete cases for all clinical risk factors included in each scenario were used in the primary analyses. Clinical risk factors in each scenario are as follows: Scenario 1) age, sex; Scenario 2) age, sex, BMI, family history of diabetes, systolic blood pressure; Scenario 3) age, sex, BMI, family history of diabetes, systolic blood pressure, random glucose; Scenario 4) age, sex, BMI, family history of diabetes, systolic blood pressure, triglycerides, total cholesterol, and HDL combined into a clinical risk score and random glucose.

|  | **Scenario 1** | **Scenario 2** | **Scenario 3** | **Scenario 4** |
| --- | --- | --- | --- | --- |
| n | 14712 | 13670 | 9867 | 7331 |
| Age, mean (SD) | 48.9 (15.7) | 48.9 (15.4) | 51.3 (14.6) | 53.9 (13.2) |
| Female, n (%) | 7965 (54.1%) | 7430 (54.4%) | 5124 (51.9%) | 3480 (47.5%) |
| Current Smokers, n (%) | 902 (6.1%) | 766 (5.2%) | 554 (3.8%) | 352 (2.4%) |
| Race |  |  |  |  |
| White, n (%) | 12697 (86.3%) | 11814 (86.4%) | 8592 (87.1%) | 6511 (88.8%) |
| Black/African American, n (%) | 525 (3.6%) | 492 (3.6%) | 373 (3.8%) | 267 (3.6%) |
| Asian, n (%) | 330 (2.2%) | 293 (2.1%) | 187 (1.9%) | 119 (1.6%) |
| Other/Unknown, n (%) | 1160 (7.9%) | 1071 (7.8%) | 715 (7.2%) | 434 (5.9%) |
| Ethnicity |  |  |  |  |
| Hispanic, n (%) | 227 (1.5%) | 185 (1.4%) | 123 (1.2%) | 80 (1.1%) |
| Non-hispanic, n (%) | 12407 (84.3%) | 11502 (84.1%) | 8294 (84.1%) | 6139 (83.7%) |
| Other/Unknown, n (%) | 2078 (14.4%) | 1983 (14.5%) | 1450 (14.7%) | 1112 (15.2%) |
| Genetic Ancestry |  |  |  |  |
| EUR, n (%) | 12508 (85.0%) | 11639 (85.1%) | 8450 (85.6%) | 6406 (87.4%) |
| AFR, n (%) | 520 (3.5%) | 489 (3.6%) | 377 (3.8%) | 269 (3.7%) |
| AMR, n (%) | 1003 (6.8%) | 924 (6.8%) | 627 (6.4%) | 357 (4.9%) |
| ASIAN, n (%) | 357 (2.4%) | 316 (2.3%) | 202 (2.0%) | 134 (1.8%) |
| Other, n (%) | 324 (2.2%) | 302 (2.2%) | 211 (2.1%) | 165 (2.3%) |
| Highest Educational Attainment |  |  |  |  |
| High School, n (%) | 3364 (22.9%) | 3113 (22.8%) | 2357 (23.9%) | 1683 (23.0%) |
| Undergraduate, n (%) | 6378 (43.4%) | 5956 (43.6%) | 4134 (41.9%) | 3045 (41.5%) |
| Graduate, n (%) | 2195 (14.9%) | 2088 (15.3%) | 1458 (14.8%) | 1106 (15.1%) |
| Other/Unknown, n (%) | 2775 (18.9%) | 2513 (18.4%) | 1918 (19.4%) | 1497 (20.4%) |
| Diagnoses at baseline |  |  |  |  |
| Coronary Artery Disease, n (%) | 713 (4.8%) | 627 (4.6%) | 573 (5.8%) | 531 (7.2%) |
| Chronic Kidney Disease, n (%) | 376 (2.6%) | 321 (2.3%) | 301 (3.1%) | 264 (3.6%) |
| Hypertension, n (%) | 3247 (22.1%) | 3046 (22.3%) | 2793 (28.3%) | 2410 (32.9%) |
| Chronic Heart Failure, n (%) | 207 (1.4%) | 177 (1.3%) | 162 (1.6%) | 126 (1.7%) |
| Peripheral Vascular Disease, n (%) | 136 (0.9%) | 115 (0.8%) | 103 (1.0%) | 78 (1.1%) |
| Family History of Diabetes, n (%) | 581 (3.9%) | 555 (4.1%) | 413 (4.2%) | 309 (4.2%) |
| Family History of CAD, n (%) | 4319 (29.4%) | 4109 (30.1%) | 3018 (30.6%) | 2330 (31.8%) |
| Incident Diabetes during follow-up, n (%) | 1908 (13.0%) | 1800 (13.2%) | 1487 (15.1%) | 1265 (17.3%) |
| BMI, mean (SD) |  | 28.3 (5.7) | 28.5 (5.7) | 28.7 (5.6) |
| Systolic Blood Pressure, mean (SD) |  | 125.7 (11.8) | 126.6 (11.7) | 127.6 (11.3) |
| Diastolic Blood Pressure, mean (SD) |  | 75.0 (6.7) | 74.9 (6.6) | 74.9 (6.5) |
| Random Glucose, mean (SD) |  |  | 97.2 (22.9) | 96.9 (22.0) |
| Total Cholesterol, mean (SD) |  |  |  | 189.9 (35.9) |
| HDL, mean (SD) |  |  |  | 56.5 (17.1) |
| Triglyceride, mean (SD) |  |  |  | 128.2 (86.6) |

Kaplan-Meier curves demonstrated full separation of PGS tertiles among all patients and in the subset of patients of European ancestry, who are most genetically similar to the cohorts used to derive the T2D PGS (Supplementary Fig. S1A, S1B). However, among the patients not of European ancestry, we observed poorer separation of PGS tertiles (Supplementary Fig. S1C). Adjusting T2D PGS by genetic ancestry probabilities (see methods) improved separation of PGS tertiles for this group (Supplementary Fig. S1D, S1E, and S1F), and corrected bias caused by PGS distribution differences across genetic ancestries; thus, residualized, ancestry-adjusted PGS were used in all subsequent analyses and all patients were analyzed as a single cohort.

T2D PGS was associated with incident diabetes and had similar HR in all scenarios adjusting only for PCs (Scenario 1 HR per SD of PGS: 1.67 (95% CI 1.59-1.74, p=1.5×10^-104^) (Fig. 1A, Supplementary Table S3). The association was preserved even after adjusting for clinical variables available in each of the scenarios. Predictive performance improved in every scenario when adding PGS to the clinical risk variables model (LRT, *P* < 0.001) (Fig. 1A, Supplementary Table S3). The improvement was most appreciable in scenario 1 with only age, sex, and PCs in the base model. The adjusted HR was 1.76 per SD of the PGS (95% CI 1.68-1.84; *P*=1.1×10^-124^; c-index improvement: 0.065) and the adjusted HR of the top 5% of the PGS compared to the IQR was 2.80 (95% CI 2.39-3.28; *P*=1.3×10^-37^; Supplementary Table S3, Supplementary Table S6). In scenario 4, with more clinical variables in the base model, the adjusted HR was 1.48 per SD of the PGS (95% CI 1.40-1.57; *P*=2.0×10^-39^; c-index improvement over base model: 0.01) and the adjusted HR of the PGS top 5% of the PGS compared to the IQR was 2.09 (95% CI 1.72-2.55; *P*=1.7×10^-13^; Supplementary Table S3, Supplementary Table S6). The performance of the PGS was lower among the patients of non-European ancestries (Supplementary Table S4, Supplementary Table S5) likely because the PGS were derived from summary statistics of meta-analyses GWAS performed on cohorts with an overrepresentation of European descent. We conducted two sensitivity analyses for the clinical base model in scenario 4. First, we performed multiple imputation for missing lab values (<8% missing). Second, we used the individual clinical variables without combining them in a CRS. The improvement of the c-index when adding PGS to the base clinical model was similar when using multiple imputation and when using individual clinical variables instead of the CRS (Supplementary Table S3).

**Figure 1:**
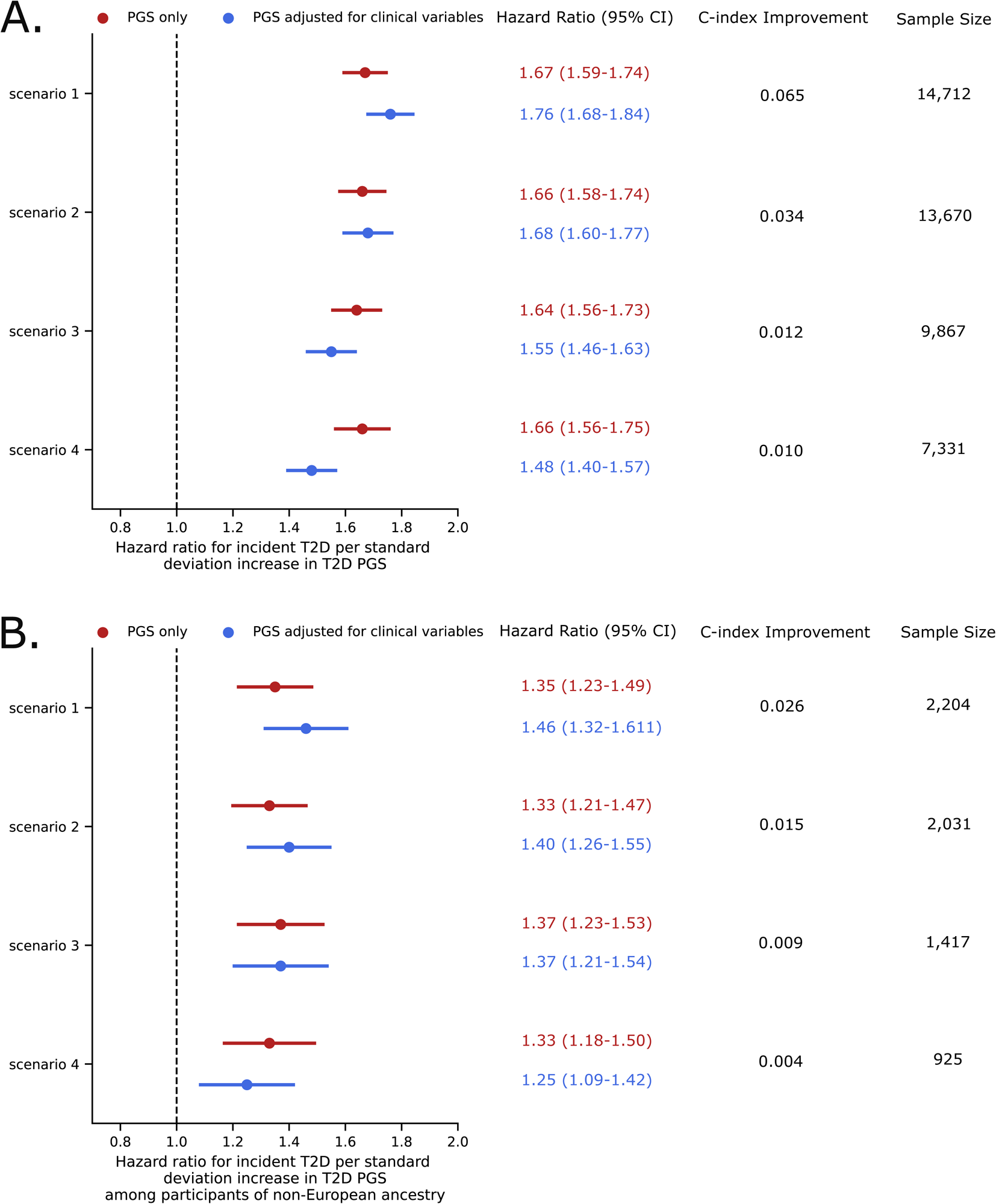
Association of T2D PGS with incident diabetes with and without adjustment for clinical variables and improvement in model performance. In all four scenarios of clinical data availability, the T2D PGS provide additional predictive information on top of clinical risk factors based on T2D PGS HR adjusted for clinical risk factors (T2D PGS adjusted HR of 1.75 in scenario 1, 1.68 in scenario 2, 1.54 in scenario 3, and 1.47 in scenario 4) and c-index improvements of including T2D PGS in clinical risk models. These benefits are largest in scenarios of minimal data availability and is true both among (A) the total cohort and (B) participants of non-European ancestry only. Clinical risk factors in each scenario are as follows: Scenario 1) age, sex; Scenario 2) age, sex, BMI, family history of diabetes, systolic blood pressure; Scenario 3) age, sex, BMI, family history of diabetes, systolic blood pressure, random glucose; Scenario 4) age, sex, BMI, family history of diabetes, systolic blood pressure, triglycerides, total cholesterol, and HDL combined into a clinical risk score and random glucose.

We found statistical interaction (*P*_interaction_ <0.05), between T2D PGS and the clinical risk factor most associated with incident diabetes in scenarios 1, 2, and 3 (i.e., age, BMI, and random glucose), and a borderline interaction between T2D CRS and PGS in scenario 4 (*P*_interaction_=0.053), motivating stratified analyses by clinical risk factors (Supplemental Table S7). The PGS estimates were larger among individuals age <40 years (HR 1.88; 95% CI 1.66-2.13; n=4,407; c-index=0.73) vs. age ≥40 years (HR 1.74; 95% CI 1.65-1.82; n=10,305; c-index=0.70) (Supplemental Table S7), among individuals with BMI <27.5 kg/m^2^ (HR 1.78, 95% CI 1.62-1.97; n=6,930; c-index=0.78) vs. BMI≥27.5 kg/m^2^ (HR 1.65, 95% CI 1.56-1.74; n=6,740; c-index=0.73) (Supplemental Table S7), among individuals with glucose <100 mg/dl (HR 1.59; 95% CI 1.46-1.74; n=6,903; c-index=0.79) vs. glucose >100 mg/dl (HR 1.48; 95% CI 1.38-1.58; n=2,964; c-index=0.77) (Supplemental Table S7), and with T2D CRS <median (HR 1.60; 95% CI 1.43-1.79; n=3,665; c-index=0.82) vs. > median (HR 1.45; 95% CI 1.35-1.55; n=3,666; c-index=0.78) (Supplemental Table S7).

In addition to hazard ratios, we also calculated each patient’s absolute risk for diabetes in each scenario. Patients aged <40 years in the highest PGS tertile had higher absolute risk [median absolute risk: 10.1% (IQR 6.9%, 15.3%)] compared to patients age ≥40 years in the lowest PGS tertile [median absolute risk: 7.2% (IQR 5.0%, 10.2%)] (Fig. 2E, Fig. 3A). Patients with BMI <27.5 kg/m^2^ in the highest PGS tertile had similar absolute risk [median absolute risk: 8.7% (IQR 4.6%, 15.7%)] to patients with BMI >27.5 kg/m^2^ in the lowest PGS tertile [median absolute risk: 8.3% (IQR 4.6%, 14.0%)] (Fig. 2F, Fig. 3B). Individuals age ≥40 years or BMI ≥27.5 kg/m^2^ in the highest PGS tertile had a greater than 25% chance of developing diabetes during the follow up period.

**Figure 2:**
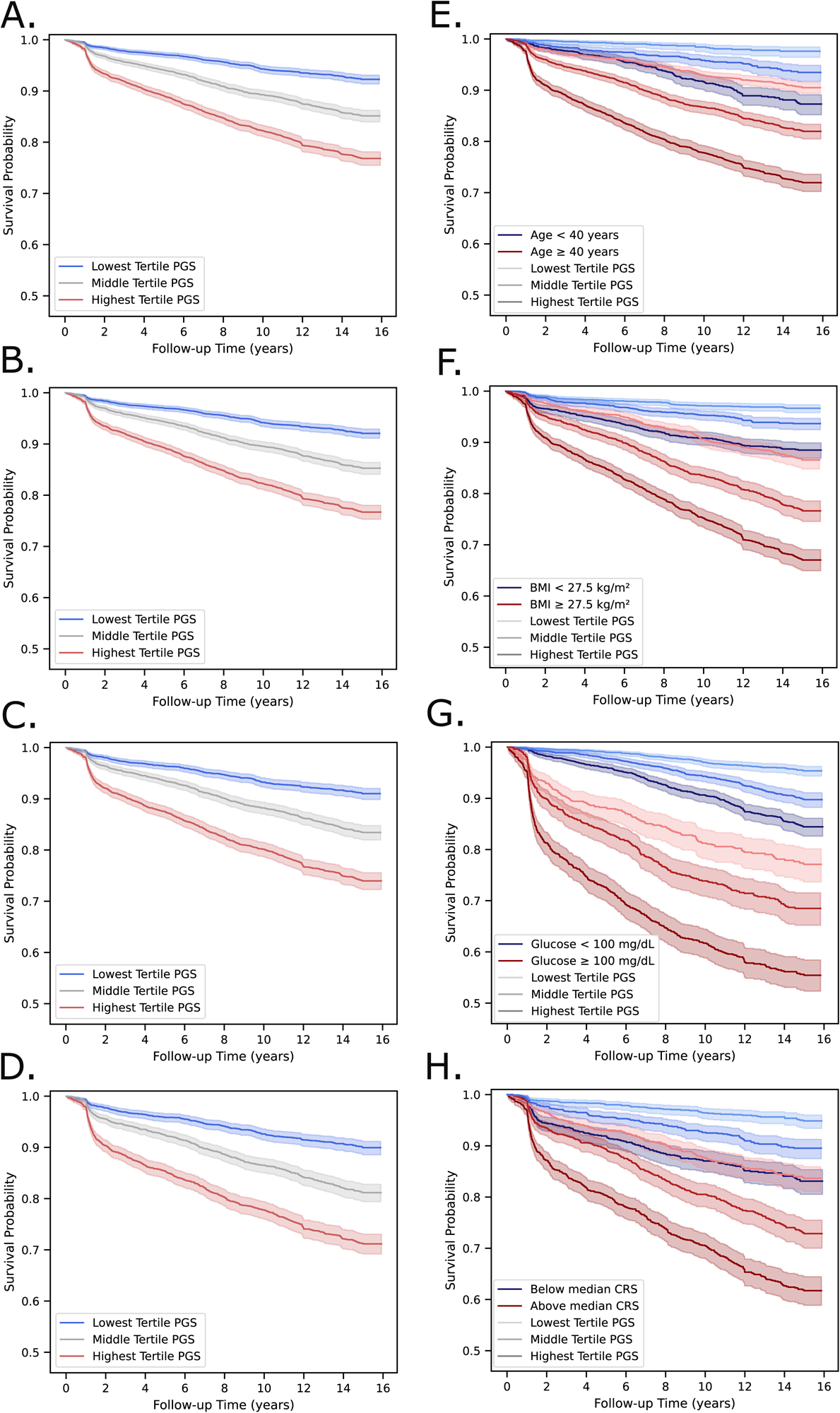
Kaplan-meier curves with and without stratification by baseline clinical risk factors. T2D PGS tertiles show strong separation of DM onset in (A) scenario 1, (B) scenario 2, (C) scenario 3, and (D) scenario 4. In each scenario, the multivariate log-rank test *P* < 0.0001. T2D PGS tertiles further stratify risk over available clinical risk factors at each scenario, including (E) an age cutoff of 40 years in scenario 1, (F) a BMI cutoff of 27.5 kg/m^2^ in scenario 2, (G) a random glucose of 100 mg/dL in scenario 3, and (H) the median T2D CRS in scenario 4. In each stratification analysis per scenario, the multivariate log-rank test *P* < 0.0001.

**Figure 3:**
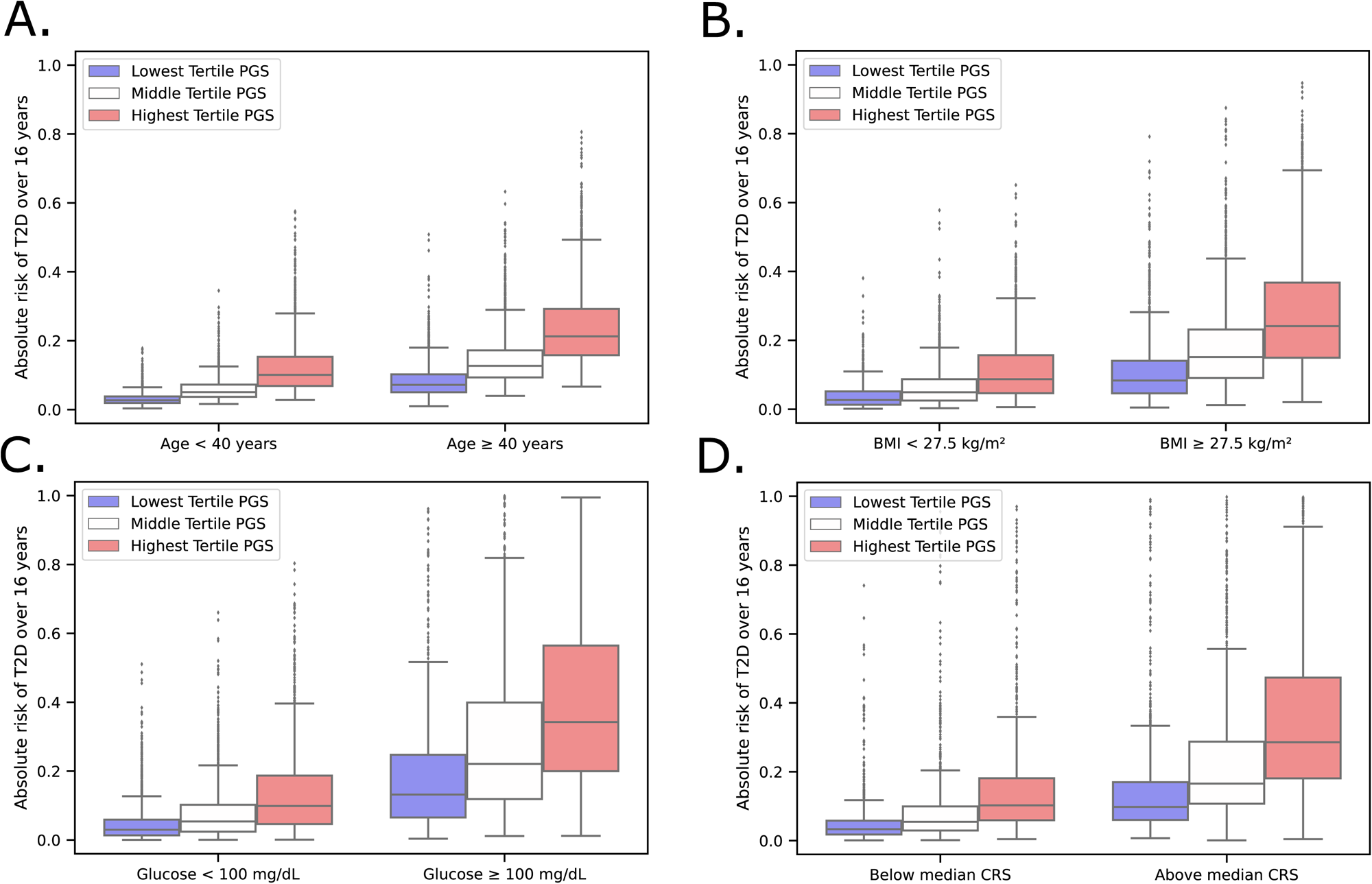
Absolute risks of developing diabetes over 16 years by T2D PGS and clinical risk factors. Patients with low clinical and low genetic risk for T2D have the lowest absolute risk for developing diabetes relative to patients with high clinical and high genetic risk. Patients with high clinical risk and low genetic risk also have similar absolute risk for developing diabetes compared to patients with low clinical risk and high genetic risk in (A) scenario 1, (B) scenario 2, (C) scenario 3, and (D) scenario 4.

Individuals with glucose <100 mg/dl had lower absolute risk in all three PGS tertiles [lowest PGS tertile: 3.0% median absolute risk (IQR 1.3%, 5.9%); highest PGS tertile: 9.9% median absolute risk (IQR 4.6%, 18.7%)] compared to those with glucose ≥100 mg/dL [lowest PGS tertile: 13.2% median absolute risk (IQR 6.5%, 24.7%); highest PGS tertile: 34.2% median absolute risk (IQR 20.0%, 56.4%)] (Fig. 2G, Fig. 3C). Similarly, individuals below the median CRS had similar or lower risk across all three PGS tertiles [lowest tertile: 3.3% median absolute risk (IQR 1.8%, 5.8%); highest tertile: 10.3% median absolute risk (IQR 5.9%, 18.1%)] compared to patients above the median CRS [lowest PGS tertile: 9.8% median absolute risk (IQR 6.0%, 17.0%); highest PGS tertile: 28.6% median absolute risk (IQR 18.1%, 47.3%)] (Fig. 2H, Fig. 3D).

Finally, we sought to evaluate how polygenic scores for CAD and CKD could improve the prediction of the onset of these comorbidities in patients who develop and do not develop diabetes. We constructed PGS for CAD (PGS-CAD) and PGS for CKD (PGS-CKD), then generated incidence models using the pertinent clinical variables from their respective CRS available at each scenario by diabetes status. PGS-CAD and PGS-CKD were both associated with incident disease among people with diabetes after adjusting for clinical visit variables (PGS-CAD per SD: HR 1.23; 95% CI 1.12, 1.34; *P*=6.4×10^-6^; PGS-CKD per SD: HR 1.29; 95% CI 1.17, 1.43; *P*=7.9×10^-7^) and when additional adjusting for clinical visit plus lab variables (PGS-CAD per SD: HR 1.13; 95% CI 1.02, 1.25; *P*=1.5×10^-2^; PGS-CKD per SD: HR 1.33; 95% CI 1.18, 1.50; *P*=4.5×10^-6^). Results were similar among patients who remained diabetes-free throughout the study period. C-index improvement, though significant, were ≤1% over the clinical base models (Supplementary Table S8).

## Conclusions

The incorporation of genetic information in the clinician’s toolbox promises to improve disease risk prediction and inform individualized health recommendations, preventive strategies, and medical decisions (29). Yet, genetic liability, modeled by PGS, has been criticized as having minimal contributions to risk prediction beyond traditional clinical risk factors (12,30). We designed four hypothetical scenarios to mimic real-world clinical settings that varied in clinical information availability, and tested whether PGS could add value to longitudinal risk prediction of diabetes in an ancestrally and medically diverse patient population with 16 years of follow-up time in a PCP network. We showed that PGS significantly improved model performance over available clinical risk factors in all scenarios, with the largest improvements in scenarios with minimal clinical risk factors available. In settings where only age and sex were considered, patients in the top 5^th^ percentile of the T2D PGS had an almost threefold risk of developing incident T2D compared to those in the IQR. In contrast when additionally considering BMI, systolic blood pressure, family history of diabetes, triglycerides, total cholesterol, HDL, and random glucose, those in the top ventile had double the risk relative to the IQR of the T2D PGS for developing incident diabetes.

We detected T2D PGS interactions with age, BMI, and random glucose, and CRS, suggesting that genetic effects were larger in younger and leaner individuals with normoglycemia that have yet to accumulate diabetes-related comorbidities(31). The probability of developing diabetes during follow-up in patients with low baseline clinical risk but high genetic risk was similar to those with high baseline clinical risk.

Additionally, patients with a high glucose or CRS and high PGS had a 20% higher chance of developing T2D compared to those with a low PGS. An efficient use of healthcare resources could include targeted screening and preventive interventions in patients with both high clinical and genetic risk. Furthermore, as many clinical risk factors, unlike genetics, are modifiable, patients identified to have high clinical risk despite low genetic risk are probably more likely to benefit from risk factor modifications. Our work is closely aligned with the mission of the Electronic Medical Records and Genomics (eMERGE) network(32), which aims to improve the combined use of both genetic and EHR data in informing healthcare decisions. We approached the issue of incorporating PGS and EHR data by considering the degree of data sparsity at the time of the initial encounter in the healthcare system, a common scenario in real-world clinical practice yet understudied in previous work. Patients lacking clinical data in the EHR are likely to be younger with few or no established comorbidities, or less engaged in preventive care. As genetic effects were found to be largest when only demographics were considered, PGS could be a powerful tool for encouraging individuals with high genetic risk to undergo a clinical risk assessment.

Our study also evaluated prediction of CAD and CKD, two diabetes-related complications that contribute largely to the morbidity and mortality of patients with diabetes. We found prediction of incident CAD and CKD significantly improved upon adding their respective PGS to clinical base models, though the improvements were smaller than what was observed for T2D in our primary analysis. Future studies on the genetic underpinnings of diabetes-related complications may enhance our understanding on how best to leverage genetic information in the prevention of diabetes-related complications.

Our study has limitations. Since the PCP network was not a closed system, incident cases could be missed when patients leave the network. Capture of these cases would still be possible if they had returned to the network during the follow up time, though the date of capture could be after the actual date of diagnosis, extending the observation time, and biasing our results towards the null. Conversely, we recognize possible contamination by type 1 diabetes being captured by the T2D algorithm, though an overwhelming majority of adult-onset diabetes is type 2. As waist-to-hip ratio and fasting status for glucose were not captured in EHR, we were unable to include these variables in the T2D CRS. Nevertheless, our study more closely reflects the accuracy and granularity of clinical information captured by EHR. The T2D CRS was derived from a non-Hispanic White cohort, which may be less accurate in racially/ethnically diverse populations. While we successfully modeled genetic risk in an ancestrally diverse cohort primary care network, most participants were of European ancestry and the PGS performed worse in patients with less than 50% European genetic ancestry. Despite this, we observed the same trend of improved predictive performance when PGS was added to base models composed of only traditional risk factors among patients that were not of European ancestry, indicating that study findings are likely still applicable to more racially and ethnically diverse US-based healthcare systems with longitudinal care. We also acknowledge that patients with existing genetic data in the biobank may not be fully representative of the patients in the network, highlighting the importance of ensuring that access to genetic information is equitably distributed throughout a healthcare system to avoid exacerbating disparities of care of minoritized or marginalized groups that are already disproportionately affected by diabetes. We recognize that improvements to the transferability of PGS and replication in other healthcare systems is necessary to properly evaluate the clinical utility of PGS among diverse populations.

With the growing literature on the predictive accuracy of PGS in diverse populations and the increasing availability of genetic information in healthcare systems, it is becoming crucial to identify when and how PGS can be used in clinical settings. We considered a range of scenarios that varied in clinical information availability to evaluate the role of polygenic risk in diabetes prediction in primary care. The utility of PGS for the purpose of identifying high-risk individuals was greatest among those with sparse clinical data and those that were younger, leaner, had few or no established cardiometabolic comorbidities, and may be perceived to have low clinical risk following a clinical evaluation. Considering genetic risk in healthcare systems provides an additional opportunity to engage high-risk patients in preventive strategies and deploy precision medicine approaches in diabetes care.

## Acknowledgements

### Funding and Assistance

J.M.M. and R.M. is supported by American Diabetes Association Innovative and Clinical Translational Award 1-19-ICTS-068, American Diabetes Association grant #11-22-ICTSPM-16 and by NHGRI U01HG011723. A.L. is supported by grant 2020096 from the Doris Duke Foundation and the American Diabetes Association Grant 7-22-ICTSPM-23. J.B.M. is supported by NIDDK U01 DK078616 and R01 HL151855.

### Author Contributions and Guarantor Statement

R.M., P.L., B.P., J.M.M., and A.L. compiled and researched data. R.M. and A.L. contributed to the discussion and wrote the manuscript. J.C.F. and J.B.M. reviewed and edited the manuscript. All authors approved the final version of the manuscript.

J.M.M. and A.L. are the guarantors of this work and, as such, had full access to all the data in the study and take responsibility for the integrity of the data and the accuracy of the data analysis.

### Prior Presentation

This work was presented at the 2022 American Society of Human Genetics Conference Poster Symposium in Los Angeles.

## Data Availability

Datasets analyzed in this study are not publicly available but on reasonable request may be made available from the corresponding author after detailing any restrictions that may apply to preserve patient confidentiality.

