## Supplemental Materials for "Polygenic Scores for Longitudinal Prediction of Incident Type 2 Diabetes in an Ancestrally and Medically Diverse Primary Care Network"

**Supplementary Materials**

**Supplementary Table S1: Administrative and ICD code based algorithms**

| **Disease** | **Criteria** | **Test Accuracies** |
| --- | --- | --- |
| **Diabetes** | Inclusion: MGH encounter AND ICD-9 code for diabetes AND Problem list term for diabetes  OR HbA1C >6.5 in prior 3 years Exclusion: Problem list term not associated with diabetes |  |
|  | Inclusion problem list terms: DM, Diabetes Mellitus, Diabetic Nephropathy, Diabetic Neuropathy,  Diabetic Retinopathy, Diabetic foot care, Diabetic foot infection, Diabetes Mellitus without complication, Insulin dependent diabetes mellitus, Insulin dependent diabetes without complications, Type 1, Diabetes mellitus I, DM type I, Juvenile, DM type i, Type I DM, Diabetes Mellitus type 1, Juvenile onset diabetes mellitus, Type 1 diabetes mellitus uncontrolled, Type 1 diabetes mellitus well controlled, DM1, Adult onset, AODM, Type 2, DM type ii, Diabetes mellitus ii, DMii, Type 2 DM, Type ii DM, Diabetes mellitus type 2, noninsulin dependent diabetes mellitus, Type 2 diabetes mellitus uncontrolled, Type 2 diabetes mellitus well controlled, Diabetes adult-adult onset, NIDDM. | Algorithm was developed as part of the MGH patient care operations improvement project, and internally validated with chart review on 260 patients Sens = 0.99 Spec = 0.93 PPV = 0.96 NPV = 0.99 |
|  | Inclusion ICD-9 codes: 250.21, 250.23, 250.11, 250.13, 250.61, 250.63, 250.51, 250.53, 250.31,  250.33, 250.81, 250.83, 250.71, 250.73, 250.41, 250.43, 250.91, 250.93, 250.01, 250.03, 250.20, 250.22, 250.10, 250.12, 250.60, 250.62, 250.50, 250.52, 250.30, 250.32, 250.80, 250.82, 250.70, 250.72, 250.40, 250.42, 250.90, 250.92, 250.00, 250.02 |  |
|  | Exclusion problem list terms: negative, neg, r/o, risk, rule out, no, consider diabetes, borderline, gestational, maternal, pregnancy, glucose impai, impaired glucose, insipidus, pre diabetes, prediab, family history, PCOS, Polycystic Ovary Syndrome |  |
| **Coronary artery disease (CAD)** | Inclusion: Problem or procedure list term for CAD OR CPT4 procedure code for CAD intervention in prior 3 years |  |
|  | Exclusion: Patients who died during the assessment period OR no problem/procedure list terms/procedure codes for CAD |  |
|  | Inclusion problem list terms: CORONARY ARTERY DISEASE, S/P CORONARY ARTERY DISEASE, H/O CORONARY ARTERY DISEASE, CORONARY ARTERIOSCLEROSIS, MYOCARDIAL INFARCTION , S/P MYOCARDIAL INFARCTION, MYOCARDIAL INFARCT, ANGINA PECTORIS, ANGINA CARDIAC BYPASS GRAFT SURGERY, S/P CARDIAC BYPASS GRAFT SURGERY, H/O CARDIAC BYPASS GRAFT SURGERY, ISCHEMIC HEART DISEASE, ISCHEMIC HRT DIS, CORONARY ARTERY BYPASS GRAFT, S/P CORONARY ARTERY BYPASS GRAFT, CAD, CAD EQUIVALENT, CORONARY STENT, S/P CORONARY STENT, STENT IN THE LAD, CORONARY HEART DISEASE, ARTERIOSCLEROTIC HEART DISEASE, ATHEROSCLEROTIC CORONARY VASCULAR DISEASE, MI, ACUTE MI, S/P MI STEMI, S/P NSTEMI, 2 CARDIAC VESSEL BYPASSES ? LAD, CARDIAC CATH STENT, S/P CARDIAC STENTS, CABG, CORONARY ARTERIAL DISEASE, ISHEMIC CARDIOMYOPATHY, ISCHEMIC CMP, MILD VENTRICULAR POSTERIOR ISCHEMIA, S/P BALLON ANGIOPLASTY. | Algorithm was developed as part of the MGH patient care operations improvement project, and internally validated with chart review on 325 patients Sens = 0.96 Spec = 0. 99 PPV = 0.99 NPV = 0.95 |
|  | Inclusion procedure list terms: CORONARY ARTERY BYPASS GRAFT, CARDIAC BYPASS GRAFT SURGERY, PERCUTANEOUS TRANSLUMINAL CORONARY ANGIOPLASTY, PLACEMENT OF STENT IN CORONARY ARTERY CORONARY STENT, ANGIOPLASTY OF LAD, CARDIAC CATHETERIZATION W/STENT CARDIAC STENT |  |
|  | Inclusion CPT procedure codes: 33510, 33511, 33512, 33513, 33514, 33516, 33517, 33518, 33519, 33521, 33522, 33523, 33530, 33533, 33534, 33535, 33536, 92980, 92981, 92982, 92984, 92975, 92977 |  |
| **Hypertension** | Inclusion: Two ICD-9 codes for hypertension or One Problem list term for hypertension in prior 3 years |  |
|  | Exclusion: End stage renal disease, receiving dialysis, prior kidney transplant or pregnancy-related hypertension |  |
|  | Inclusion problem list terms: Hypertension Hypertensive cardiovascular disease Chronic hypertension Hypertensive heart disease Benign hypertension Benign essential hypertension Malignant essential hypertension Malignant hypertension Essential hypertension Finding of increased Blood pressure Hypertensive disorder Systolic hypertension Labile hypertension Hypertensive crisis White Coat Hypertension Secondary hypertension Malignant secondary hypertension Renovascular Hypertension | Algorithm was developed as part of the MGH patient care operations improvement project, and internally validated with chart review on 325 patients. Sens = 0.95 Spec = 0.94 PPV = 0.97 NPV = 0.89 |
|  | Inclusion ICD-9 codes: 997.91, 401.0, 401.1, 401.9, 402.01, 402.00, 402.91, 402.90, 403.01, 403.00, 404.01, 404.03, 404.91, 404.93, 404.92, 404.11, 404.13, 404.12, 404.10, 402.11, 402.10, 403.11, 403.10: |  |
|  | Exclusion codes and terms: Pregnancy induced hypertension, Gestational Hypertension, End Stage Renal disease, Hemodialysis, Dialysis, Renal Transplant, S/P Renal Transplant, Renal Transplantation, S/P Renal Transplantation, Kidney Transplant, S/P Kidney Transplant, Transplant of Kidney, Kidney Transplantation, S/P Kidney Transplantation, ICD-9 codes 796.2, 585.6, CPT codes 36800, 36810, 36815, 36818, 36819, 36820, 36821, 36831, 36832, 36833, 50300, 50320, 50340, 50360, 50365, 50370, 50380, 90935, 90937 |  |
| **Cerebrovascular disease (CVD)** | Inclusion: Problem/procedure list term for CVD OR CPT4 procedure code for CVD intervention in prior 3 years |  |
|  | Exclusion: patients who died during the assessment period OR no problem/procedure list terms/procedure codes for CVD |  |
|  | Inclusion problem list terms: STROKE, S/P STROKE, H/O STROKE, CVA, H/O CVA, S/P CVA, TIA, H/O TIA, HX OF TRANSIENT ISCHEMIC ATTACK, S/P TIA, TRANSIENT CEREBRAL ISCHEMIA, CAROTID STENOSIS. H/O CAROTID ARTERY STENOSIS, LACUNAR CVA, LACUNAR INFARCT, LACUNAR STROKE, MCA STENT, CAROTID ENDARTERECTOMY, S/P RT CEA, S/P LEFT CEA, H/O CEREBELLAR INFARCT, HX OF CEREBELLAR INFARCT, CEREBROVASCULAR ACCIDENT, H/O CEREBROVASCULAR ACCIDENT, CEREBROVASCULAR DISEASE, H/O CEREBROVASCULAR DISEASE, INTRACRANIAL ATHEROSCLEROSIS CAROTID OCCLUSION, H/O CAROTID OCCLUSION | Algorithm was developed as part of the MGH patient care operations improvement project, and internally validated with chart review on 325 patients. Sens = 0.95 Spec = 0.99 PPV = 0.99 NPV = 0.93 |
|  | Inclusion procedure list terms: CAROTID ENDARTERECTOMY, S/P CAROTID ENDARTERECTOMY, BILATERAL CAROTID ENDARTERECTOMY, H/O CAROTID ENDARTERECTOMY, CAROTID ARTERY STENT, CAROTID STENOSIS RIGHT STENT |  |
|  | Inclusion CPT Procedure codes: 35301, 33891, 35626, 35606, 35642, 35601, 35526, 35526, 35510, 35509, 35506, 35508, 35501, 35001, 34001, 35390, 35301, 35301, 37215, 37216, 61630, 61635 |  |
| **Peripheral vascular diseae (PVD)** | Inclusion: Problem/procedure list term for PVD or CPT4 procedure code for PVD intervention |  |
|  | Exclusion: patients who died during the assessment period OR no problem/procedure list terms/procedure codes for PVD |  |
|  | Inclusion problem list terms: PERIPHERAL VASCULAR DISEASE, H/O PERIPHERAL VASCULAR DISEASE, ABDOMINAL AORTIC ANEURYSM, S/P ABDOMINAL AORTIC ANEURYSM, H/O ABDOMINAL AORTIC ANEURYSM, ABDOMINAL AORTIC ANEURYSM REPAIR, AORTIC ANEURYSM, AORTIC ARCH ANEURYSM, ASCENDING AORTIC ANEURYSM, ANEURYSM OF THORACIC AORTA, ANEURYSM OF THORACIC AORTA, ASCENDING THORACIC AORTIC ANEURYSM, PAD, PERIPHERAL ARTERIAL DISEASE, PERIPHERAL ARTERIAL OCCLUSIVE DISEASE, RENAL ARTERY STENOSIS, MILD RENAL ARTERY STENOSIS, ISCHEMIC EXTREMITY, PERIPHERAL ISCHEMIA, OCCLUDED RIGHT FEM POP BYPASS, S/P AORTO BIFEMORAL BYPASS, FEM POP BYPASS, S/P FEMORAL POPLITEAL BYPASS, ANEURYSM, ILIAC, AORTIC STENOSIS, ARTERIOSCLEROTIC VASCULAR DISEASE, BILATERAL ILIAC ARTERY STENOSIS, ILIAC ARTERY ANEURYSM, S/P ILIAC PTCA/STENT, STENT IN BILAT EXERTENAL ILIAC ARTERIES | Algorithm was developed as part of the MGH patient care operations improvement project, and internally validated with chart review on 325 patients. Sens = 0.92 Spec = 0.99 PPV = 0.99 NPV = 0.90 |
|  | Inclusion procedure list terms: FEMORAL POPLITEAL BYPASS, FEMORAL POPLITEAL ARTERY BYPASS GRAFT, AORTOFEM BYPASS, AAA REPAIR, ABDOMINAL AORTIC ANEURYSM REPAIR, ABDOMINAL AORTIC ANEURYSM STENT, ILIAC STENT, ABDOMINAL AORTIC ANEURYSM STENT, S/P ABDOMINAL AORTIC ANEURYSM STENT, AORTO BIFEMORAL BYPASS GRAFT, AORTO FEM BYPASS, AORTOFEMORAL BYPASS, AORTOFEMORAL BYPASS GRAFT, AORTOILIAC ANGIOPLASTY, BILATERAL FEMORAL POPLITEAL BYPASS, ENDOVASCULAAR REAPIR OF ABDOMINAL AORTIC, FEMORAL ARTERY STENT, FEMORAL POPLITEAL ARTERY BYPASS GRAFT, FEMOROPOPLITEAL BYPASS, LEFT FEMORAL ARTERY STENT, RENAL ARTERY STENOSIS STENT, RENAL ARTERY STENT, RENAL ARTERY STENTS, S/P ABDOMINAL AORTA ENDOGRAFT REPAIR, ILIAC ARTERY STENT GRAFT, ILIAC ARTERY STENTING, L FEMORAL, ENDARTERECTOMY, THORACIC AORTIC ANEURYSM REPAIR, THORACOABDOMINAL ANEURYSM REPAIR |  |
|  | Inclusion CPT Procedure codes: 35566, 35141, 35142, 35131, 35132, 34201, 34151, 34808, 34820, 34151, 35450, 35471, 35454, 35473, 35452, 35472, 35480, 35490, 35646, 35647, 35651, 35621, 35654, 35654, 35666, 35661, 35656, 35665, 35540, 35638, 35637, 35663, 35538, 35537, 35563, 35081, 35102, 34151, 34823, 34825, 33877, 35549, 35548, 35450 |  |
| **Congestive Heart Failure (CHF)** | Inclusion: Adult patients (18 and older) with the following: 1) One inpatient primary discharge diagnosis of CHF OR 2) For patients without inpatient code for CHF, first instances of two outpatient visits with problem list terms or billing diagnoses for CHF in any combination |  |
|  | Exclusions: Any problem list term/CPT code OR any two ICD-9 diagnosis codes |  |
|  | Inclusion problem list terms: CORONARY ARTERY DISEASE, S/P CORONARY ARTERY DISEASE, H/O CORONARY ARTERY DISEASE, CORONARY ARTERIOSCLEROSIS, MYOCARDIAL INFARCTION , S/P MYOCARDIAL INFARCTION, MYOCARDIAL INFARCT, ANGINA PECTORIS, ANGINA CARDIAC BYPASS GRAFT | Algorithm was developed as part of the MGH patient care operations improvement project, and internally validated with chart review on 358 patients. Sens = 0.99 Spec = 0.85 PPV = 0.86 NPV = 0.99 |
|  | Inclusion ICD-9 codes: 428.0, 428.1, 428.20, 428.21, 428.22, 428.23, 428.30, 428.31, 428.32, 428.33,  428.40, 428.41, 428.42, 428.43, 428.9, 398.91, 404.11, 404.13, 402.11, 404.01, 404.03, 402.01, 402.91, 404.91, 404.93, 425.11, 425.4, 425.5, 425.7, 425.8, 425.9 |  |
|  | Exclusion problem list terms: Myocarditis, Viral cardiomyopathy, Heart transplant, S/P Heart  transplant |  |
|  | Exclusion ICD-9 codes: 391.2, 398.0, 422.0, 422.90, 422.91, 422.92, 429.0, 074.23, 032.82, 036.43,  130.3, 422.99, 093.82, 422.93, 422.90, V42.1, 996.83, 33945, 33935, 33944, 33933 |  |
|  | Exclusion CPT Procedure codes: 33945, 33935, 33944, 33933 |  |
| **Chronic Kidney Disease (CKD) and  endstage CKD** | Inclusion: Adult patients (18 to 84) with Average eGFR <55, last eGFR <=59, more than one eGFR drawn 2009-2010 |  |
|  | Exclusion: End Stage Renal Disease, Kidney Transplant, Avg eGFR <15 |  |
|  | Exclusion problem list terms: End Stage Renal disease, Renal Transplant, S/P Renal Transplant, Renal Transplantation, S/P Renal Transplantation, Kidney Transplant, S/P Kidney Transplant, Transplant of Kidney, Kidney Transplantation, S/P Kidney Transplantation | Algorithm was developed as part of the MGH patient care operations improvement project patients. Spec = 0.90 PPV = 0.90 |
|  | Exclusion ICD codes: 585.6, N18.6, I12.0, I13.2, I13.11, N18.5 |  |
|  | Exclusion CPT Procedure codes: 50300, 50320, 50360, 50365, 50370, 50380 |  |
| **Proteinuria** | Inclusion ICD codes: R80 |  |

**Supplemental Table S2: Patient characteristics of both Mass General Brigham Biobank (MGBB) and Mass General Hospital (MGH) Primary Care Physician (PCP) network**

|  | **MGBB + PCP network** | **PCP network only** | **MGBB NOT in PCP network with multiple encounters** | **MGBB NOT in PCP network without multiple encounters** |
| --- | --- | --- | --- | --- |
| **n** | 15355 | 269247 | 26454 | 11653 |
| **Age as of 2020, mean (SD)** | 61.0 (16.3) | 55.4 (18.4) | 56.6 (17.2) | 55.3 (18.8) |
| **Female, n (%)** | 8255 (53.8%) | 151268 (56.2%) | 14917 (56.4%) | 6145 (52.7%) |
| **Current Smokers, n (%)** | 954 (6.2%) | 37693 (14.0%) | 1756 (6.6%) | 577 (5.0%) |
| **Race** |  |  |  |  |
| White, n (%) | 13171 (85.8%) | 198887 (73.9%) | 22488 (85.0%) | 9547 (81.9%) |
| Black/African American, n (%) | 577 (3.8%) | 15780 (5.9%) | 1429 (5.4%) | 585 (5.0%) |
| Asian, n (%) | 342 (2.2%) | 17794 (6.6%) | 661 (2.5%) | 317 (2.7%) |
| Other/Unknown, n (%) | 1265 (8.2%) | 36786 (13.7%) | 1876 (7.1%) | 1204 (10.3%) |
| **Ethnicity** |  |  |  |  |
| Hispanic, n (%) | 240 (1.6%) | 25911 (9.6%) | 451 (1.7%) | 246 (2.1%) |
| Non-hispanic, n (%) | 12961 (84.4%) | 152154 (56.5%) | 22908 (86.6%) | 9773 (83.9%) |
| Other/Unknown, n (%) | 2154 (14.0%) | 91182 (33.9%) | 3095 (11.7%) | 1634 (14.0%) |
| **Highest Educational Attainment** |  |  |  |  |
| High School, n (%) | 3631 (23.6%) | 46176 (17.2%) | 7575 (28.6%) | 3399 (29.2%) |
| Undergraduate, n (%) | 6564 (42.7%) | 93833 (34.9%) | 11672 (44.1%) | 4464 (38.3%) |
| Graduate, n (%) | 2273 (14.8%) | 14323 (5.3%) | 3549 (13.4%) | 1196 (10.3%) |
| Other/Unknown, n (%) | 2887 (18.8%) | 114915 (42.7%) | 3658 (13.8%) | 2594 (22.3%) |
| **Diabetes Prevalence, n (%)** | 2551 (16.6%) | 23022 (8.6%) | 161 (0.6%) | 8 (0.1%) |

**Supplemental Table S3: Association of polygenic scores as a continuous variable with incidence diabetes adjusting for clinical variables in the base model and change in model performance in the entire population.** Longitudinal models were constructed with either clinical variables included in each scenario, polygenic scores (PGS) only, or both the clinical variables and PGS in a combined model. Clinical risk factors in each scenario are as follows: Scenario 1) age, sex; Scenario 2) age, sex, BMI, family history of diabetes, systolic blood pressure; Scenario 3) age, sex, BMI, family history of diabetes, systolic blood pressure, random glucose; Scenario 4) age, sex, BMI, family history of diabetes, systolic blood pressure, triglycerides, total cholesterol, and HDL combined into a clinical risk score (CRS) and random glucose. Two sensitivity analyses for Scenario 4 were run: Complete-case analysis (CCA), where variables were not combined into a CRS and only participants without any missing data were included; Multiple Imputation (MI), where variables were not combined into a CRS and missing lab values from participants without full data were imputed. We reported the concordance index (C-index), Hazard Ratio (HR) for the PGS or CRS per standard deviation depending on if they are included in the model, and the log-likelihood ratio test (LRT) p-value from comparing the difference in performance between the Combined Clinical and PGS Model with the Clinical Variables Only Model.

|  | **Scenario 1** | **Scenario 2** | **Scenario 3** | **Scenario 4** | **Scenario 4 - CCA** | **Scenario 4 - MI** |
| --- | --- | --- | --- | --- | --- | --- |
| **n** | 14712 | 13670 | 9867 | 7331 | 7331 | 8390 |
| **Clinical Variables Only Model** |  |  |  |  |  |  |
| C-index | 0.663 | 0.749 | 0.817 | 0.806 | 0.832 | 0.834 |
| CRS HR (CI) (p-val) |  |  |  | 1.75 (1.63-1.88) (3.6e-54) |  |  |
| **PGS Only Model** |  |  |  |  |  |  |
| C-index | 0.667 | 0.667 | 0.663 | 0.667 | 0.667 | 0.668 |
| PGS HR (CI) (p-val) | 1.67 (1.59-1.74) (1.5E-104) | 1.66 (1.58-1.74) (1.0E-96) | 1.64 (1.56-1.73) (3.1E-77) | 1.66 (1.56-1.75) (5.6E-67) | 1.66 (1.56-1.75) (5.6E-67) | 1.66 (1.57-1.76) (3.6E-72) |
| **Combined Clinical and PGS Model** |  |  |  |  |  |  |
| C-index | 0.728 | 0.783 | 0.829 | 0.816 | 0.841 | 0.843 |
| CRS HR (CI) (p-val) |  |  |  | 1.71 (1.60-1.84) (6.5E-50) |  |  |
| PGS HR (CI) (p-val) | 1.76 (1.68-1.84) (1.1E-124) | 1.68 (1.6-1.77) (4.1E-98) | 1.55 (1.46-1.63) (2.4E-55) | 1.48 (1.40-1.57) (2.0E-39) | 1.48 (1.39-1.57) (1.4E-37) | 1.49 (1.41-1.58) (1.3E-41) |
| **C-index Improvement** | 0.065 | 0.034 | 0.012 | 0.01 | 0.009 | 0.009 |
| **LRT p-value** | 1.03E-124 | 4.52E-98 | 2.12E-55 | 1.43E-39 | 8.28E-38 | 7.40E-42 |

**Supplemental Table S4: Association of polygenic scores as a continuous variable with incidence diabetes adjusting for clinical variables in the base model and change in model performance in participants of European ancestry.** Longitudinal models among only participants of European ancestry were constructed with either clinical variables included in each scenario, polygenic scores (PGS) only, or both the clinical variables and PGS in a combined model. Clinical risk factors in each scenario are as follows: Scenario 1) age, sex; Scenario 2) age, sex, BMI, family history of diabetes, systolic blood pressure; Scenario 3) age, sex, BMI, family history of diabetes, systolic blood pressure, random glucose; Scenario 4) age, sex, BMI, family history of diabetes, systolic blood pressure, triglycerides, total cholesterol, and HDL combined into a clinical risk score (CRS) and random glucose. Two sensitivity analyses for Scenario 4 were run: Complete-case analysis (CCA), where variables were not combined into a CRS and only participants without any missing data were included; Multiple Imputation (MI), where variables were not combined into a CRS and missing lab values from participants without full data were imputed. We reported the concordance index (C-index), Hazard Ratio (HR) for the PGS or CRS per standard deviation depending on if they are included in the model, and the log-likelihood ratio test (LRT) p-value from comparing the difference in performance between the Combined Clinical and PGS Model with the Clinical Variables Only Model.

|  | Scenario 1 | Scenario 2 | Scenario 3 | Scenario 4 | Scenario 4 - CCA | Scenario 4 - MI |
| --- | --- | --- | --- | --- | --- | --- |
| **n** | 12508 | 11639 | 8450 | 6406 | 6406 | 7327 |
| **Clinical Variables Only Model** |  |  |  |  |  |  |
| C-index | 0.644 | 0.746 | 0.818 | 0.807 | 0.832 | 0.833 |
| CRS HR (CI) (p-val) |  |  |  | 1.82 (1.69-1.97) (2.5E-50) |  |  |
| **PGS Only Model** |  |  |  |  |  |  |
| C-index | 0.669 | 0.668 | 0.662 | 0.665 | 0.665 | 0.665 |
| PGS HR (CI) (p-val) | 1.84 (1.74-1.94) (1.4E-108) | 1.83 (1.73-1.94) (3.4E-102) | 1.79 (1.69-1.9) (3.1E-79) | 1.82 (1.71-1.95) (2.3E-71) | 1.82 (1.71-1.95) (2.3E-71) | 1.82 (1.71-1.94) (1.3E-76) |
| **Combined Clinical and PGS Model** |  |  |  |  |  |  |
| C-index | 0.726 | 0.787 | 0.832 | 0.82 | 0.843 | 0.845 |
| CRS HR (CI) (p-val) |  |  |  | 1.78 (1.64-1.92) (9.1E-46) |  |  |
| PGS HR (CI) (p-val) | 1.93 (1.83-2.04) (2.7E-125) | 1.83 (1.73-1.94) (7.4E-98) | 1.61 (1.52-1.72) (3.2E-50) | 1.55 (1.45-1.66) (5.3E-38) | 1.54 (1.44-1.65) (1.8E-35) | 1.55 (1.45-1.66) (4.6E-39) |
| **C-index Improvement** | 0.082 | 0.041 | 0.014 | 0.013 | 0.011 | 0.012 |
| **LRT p-value** | 8.44E-126 | 5.99E-98 | 2.34E-50 | 3.42E-38 | 7.89E-36 | 1.84E-39 |

**Supplemental Table S5: Association of polygenic scores as a continuous variable with incidence diabetes adjusting for clinical variables in the base model and change in model performance in participants not of European ancestry.** Longitudinal models among only participants not of European ancestry were constructed with either clinical variables included in each scenario, polygenic scores (PGS) only, or both the clinical variables and PGS in a combined model. Clinical risk factors in each scenario are as follows: Scenario 1) age, sex; Scenario 2) age, sex, BMI, family history of diabetes, systolic blood pressure; Scenario 3) age, sex, BMI, family history of diabetes, systolic blood pressure, random glucose; Scenario 4) age, sex, BMI, family history of diabetes, systolic blood pressure, triglycerides, total cholesterol, and HDL combined into a clinical risk score (CRS) and random glucose. Two sensitivity analyses for Scenario 4 were run: Complete-case analysis (CCA), where variables were not combined into a CRS and only participants without any missing data were included; Multiple Imputation (MI), where variables were not combined into a CRS and missing lab values from participants without full data were imputed. We reported the concordance index (C-index), Hazard Ratio (HR) for the PGS or CRS per standard deviation depending on if they are included in the model, and the log-likelihood ratio test (LRT) p-value from comparing the difference in performance between the Combined Clinical and PGS Model with the Clinical Variables Only Model.

|  | **Scenario 1** | **Scenario 2** | **Scenario 3** | **Scenario 4** | **Scenario 4 - CCA** | **Scenario 4 - MI** |
| --- | --- | --- | --- | --- | --- | --- |
| **n** | 2204 | 2031 | 1417 | 925 | 925 | 1063 |
| **Clinical Variables Only Model** |  |  |  |  |  |  |
| C-index | 0.704 | 0.744 | 0.793 | 0.774 | 0.806 | 0.812 |
| CRS HR (CI) (p-val) |  |  |  | 1.45 (1.23-1.72) (9.4e-06) |  |  |
| **PGS Only Model** |  |  |  |  |  |  |
| C-index | 0.62 | 0.622 | 0.621 | 0.618 | 0.618 | 0.625 |
| PGS HR (CI) (p-val) | 1.35 (1.23-1.49) (3.8E-10) | 1.33 (1.21-1.47) (8.6E-09) | 1.37 (1.23-1.53) (1.4E-08) | 1.33 (1.18-1.50) (2.1E-06) | 1.33 (1.18-1.50) (2.1E-06) | 1.33 (1.18-1.50) (1.6E-06) |
| **Combined Clinical and PGS Model** |  |  |  |  |  |  |
| C-index | 0.73 | 0.759 | 0.802 | 0.778 | 0.81 | 0.816 |
| CRS HR (CI) (p-val) |  |  |  | 1.42 (1.20-1.67) (3.5E-05) |  |  |
| PGS HR (CI) (p-val) | 1.46 (1.32-1.61) (1.9E-14) | 1.4 (1.26-1.55) (8.0E-11) | 1.37 (1.21-1.54) (2.4E-07) | 1.25 (1.09-1.42) (8.8E-04) | 1.31 (1.14-1.49) (9.2E-05) | 1.31 (1.15-1.49) (6.1E-05) |
| **C-index Improvement** | 0.026 | 0.015 | 0.009 | 0.004 | 0.004 | 0.004 |
| **LRT p-value** | 2.05E-14 | 8.60E-11 | 2.18E-07 | 8.72E-04 | 9.09E-05 | 5.94E-05 |

**Supplemental Table S6:** **Association of polygenic scores as a categorical variable (top 5% vs interquartile range) with incidence diabetes adjusting for clinical variables in the base model and change in model performance**. Longitudinal models were constructed with either clinical variables included in each scenario, polygenic scores (PGS) only, or both the clinical variables and PGS in a combined model. PGS were converted into a categorical value differentiating participants in the top 5% of the PGS compared to those in the interquartile range (IQR) of the PGS. Clinical risk factors in each scenario are as follows: Scenario 1) age, sex; Scenario 2) age, sex, BMI, family history of diabetes, systolic blood pressure; Scenario 3) age, sex, BMI, family history of diabetes, systolic blood pressure, random glucose; Scenario 4) age, sex, BMI, family history of diabetes, systolic blood pressure, triglycerides, total cholesterol, and HDL combined into a clinical risk score (CRS) and random glucose. We reported the concordance index (C-index), Hazard Ratio (HR) for the PGS for being in the top 5% compared to the IQR of the PGS or CRS per standard deviation depending on if they are included in the model, and the log-likelihood ratio test (LRT) p-value from comparing the difference in performance between the Combined Clinical and PGS Model with the Clinical Variables Only Model.

|  | **Scenario 1** | **Scenario 2** | **Scenario 3** | **Scenario 4** |
| --- | --- | --- | --- | --- |
| **n of DM cases (n in top 5%, n in IQR)** | 1073 (199, 874) | 1002 (184,818) | 827 (152, 675) | 712 (132, 580) |
| **total n (n in top 5%, n in IQR)** | 8092 (736, 7356) | 7518 (684, 6834) | 5427 (494, 4933) | 4032 (367, 3665) |
| **Clinical Variables Only Model** |  |  |  |  |
| C-index | 0.675 | 0.752 | 0.816 | 0.801 |
| CRS HR (CI) (p-val) |  |  |  | 1.71 (1.56-1.88) (1.6e-29) |
| **PGS Only Model** |  |  |  |  |
| C-index | 0.608 | 0.607 | 0.603 | 0.608 |
| PGS HR (CI) (p-val) | 2.43 (2.08-2.85) (1.0e-28) | 2.40 (2.04-2.83) (7.1e-26) | 2.43 (2.03-2.91) (3.5e-22) | 2.58 (2.13-3.14) (7.3e-22) |
| **Combined Clinical and PGS Model** |  |  |  |  |
| C-index | 0.702 | 0.769 | 0.822 | 0.803 |
| CRS HR (CI) (p-val) |  |  |  | 1.71 (1.56-1.88) (6.6e-29) |
| PGS HR (CI) (p-val) | 2.80 (2.39-3.28) (1.3e-37) | 2.65 (2.25-3.12) (3.3e-31) | 2.40 (2.0-2.88) (4.9e-21) | 2.09 (1.72-2.55) (1.7e-13) |
| **C-index Improvement** | 0.027 | 0.017 | 0.006 | 0.002 |
| **LRT p-value** | 8.77E-31 | 6.12E-26 | 5.44E-18 | 7.09E-12 |

**Supplementary Table S7:** **Association of polygenic scores as a continuous variable with incidence diabetes adjusting for clinical variables in the base model and change in model performance by baseline clinical risk factors.** Longitudinal models were constructed with either clinical variables included in each scenario, polygenic scores (PGS) only, or both the clinical variables and PGS in a combined model. Clinical risk factors in each scenario are as follows: Scenario 1) age, sex; Scenario 2) age, sex, BMI, family history of diabetes, systolic blood pressure; Scenario 3) age, sex, BMI, family history of diabetes, systolic blood pressure, random glucose; Scenario 4) age, sex, BMI, family history of diabetes, systolic blood pressure, triglycerides, total cholesterol, and HDL combined into a clinical risk score (CRS) and random glucose. Per scenario, an interaction term between PGS and the most significantly associated clinical variable per scenario was included in the model and tested for significance. Analyses were then performed stratifying different risk cutoffs for the clinical variable which significantly interacted with PGS per scenario. The cutoffs used per scenario were: Scenario 1) Age cutoff of 40 years; Scenario 2) BMI cutoff of 27.5 kg/m2; Scenario 3) Glucose cutoff of 100 mg/dl; Scenario 4) Cutoff of Median CRS. We reported the concordance index (C-index), Hazard Ratio (HR) for the PS or CRS per standard deviation depending on if they are included in the model, and the log-likelihood ratio test (LRT) p-value from comparing the difference in performance between the Combined Clinical and PGS Model with the Clinical Variables Only Model, and the interaction p-value between the clinical variable and PGS. Analyses were run in all participants, and in subset of participants stratified by genetic ancestry.

|  | **Scenario 1** | | **Scenario 2** | | **Scenario 3** | | **Scenario 4** | |
| --- | --- | --- | --- | --- | --- | --- | --- | --- |
| **Interaction Variable (P-value)** | Age (0.028) | | BMI (6.6e-3) | | Random Glucose (0.011) | | CRS (0.053) | |
|  | Age < 40 | Age >= 40 | BMI < 27.5 | BMI >= 27.5 | Glucose < 100 | Glucose >= 100 | CRS < Median | CRS >= Median |
| **n** | 4407 | 10305 | 6930 | 6740 | 6903 | 2964 | 3665 | 3666 |
| **Clinical Variables Only Model** |  |  |  |  |  |  |  |  |
| C-index | 0.668 | 0.621 | 0.727 | 0.683 | 0.765 | 0.752 | 0.807 | 0.766 |
| CRS HR (CI) (p-val) |  |  |  |  |  |  | 2.01 (1.63-2.47) (3.6e-11) | 1.63 (1.42-1.88) (1.3e-11) |
| **PGS Only Model** |  |  |  |  |  |  |  |  |
| C-index | 0.697 | 0.682 | 0.679 | 0.647 | 0.669 | 0.633 | 0.678 | 0.646 |
| PGS HR (CI) (p-val) | 1.84 (1.62-2.08) (1.7e-21) | 1.7 (1.62-1.79) (2.6e-96) | 1.72 (1.56-1.90) (2.7e-27) | 1.57 (1.49-1.66) (1.1e-59) | 1.62 (1.49-1.76) (7.9e-28) | 1.52 (1.41-1.62) (2.0e-32) | 1.73 (1.55-1.93) (4.0e-23) | 1.57 (1.47-1.68) (1.6e-38) |
| **Combined Clinical and PGS Model** |  |  |  |  |  |  |  |  |
| C-index | 0.725 | 0.702 | 0.776 | 0.729 | 0.789 | 0.766 | 0.819 | 0.775 |
| CRS HR (CI) (p-val) |  |  |  |  |  |  | 1.97 (1.6-2.42) (1.3e-10) | 1.62 (1.40-1.86) (3.5e-11) |
| PGS HR (CI) (p-val) | 1.88 (1.66-2.13) (1.0e-22)\ | 1.74 (1.65-1.82) (2.7e-103) | 1.78 (1.62-1.97) (1.5e-30) | 1.65 (1.56-1.74) (3.2e-69) | 1.59 (1.46-1.74) (2.5e-25) | 1.48 (1.38-1.58) (1.9e-27) | 1.6 (1.43-1.79) (5.0e-16) | 1.45 (1.35-1.55) (5.2e-26) |
| **C-index Improvement** | 0.057 | 0.081 | 0.049 | 0.046 | 0.024 | 0.014 | 0.012 | 0.009 |
| **LRT p-value** | 9.36E-23 | 2.55E-103 | 2.22E-30 | 1.83E-69 | 2.55E-25 | 1.49E-27 | 4.45E-16 | 4.27E-26 |

**Supplementary Table S8: Association of polygenic scores as a continuous variable with incident coronary artery disease or chronic kidney disease adjusting for clinical variables in the base model and change in model.** Longitudinal models were constructed with either clinical variables included in each scenario, polygenic scores (PGS) only, or both the clinical variables and PGS in a combined model. Clinical risk factors for the CAD analysis included: Clinical Visit - age, sex, smoking status, and systolic blood pressure; Clinical Visit with Labs - age, sex, smoking status, systolic blood pressure, HDL, and total cholesterol combined into a clinical risk score (CRS). Clinical risk factors for the CKD analysis included: Clinical Visit - age, sex, diagnosis history, systolic blood pressure, diastolic blood pressure, weight and HTN diagnosis; Clinical Visit with Labs - variables from the "Clinical Visit" scenario combined into a CRS. Analyses were performed in patients with or without diabetes. We reported the concordance index (C-index), Hazard Ratio (HR) for the PGS or CRS per standard deviation depending on if they are included in the model, and the log-likelihood ratio test (LRT) p-value from comparing the difference in performance between the Combined Clinical and PGS Model with the Clinical Variables Only Model, and the interaction p-value between the clinical variable and PGS.

|  | **CAD** | | | | **CKD** | | | |
| --- | --- | --- | --- | --- | --- | --- | --- | --- |
|  | with Diabetes | | without Diabetes | | with Diabetes | | without Diabetes | |
|  | **Clinical Visit** | **Clinical Visit with Labs** | **Clinical Visit** | **Clinical Visit with Labs** | **Clinical Visit** | **Clinical Visit with Labs** | **Clinical Visit** | **Clinical Visit with Labs** |
| **n** | 2124 | 1589 | 11515 | 6733 | 2267 | 1343 | 11645 | 5921 |
| **Clinical Only** |  |  |  |  |  |  |  |  |
| C-index | 0.701 | 0.666 | 0.813 | 0.743 | 0.751 | 0.714 | 0.814 | 0.771 |
| CRS HR (CI) (p-val) |  | 2.05 (1.79-2.35) (1.0e-25) |  | 2.75 (2.51-3.02) (2.3e-101) |  | 4.18 (3.25-5.39) (1.1e-28) |  | 4.16 (3.53-4.90) (1.8e-65) |
| **PGS Only Model** |  |  |  |  |  |  |  |  |
| C-index | 0.574 | 0.551 | 0.589 | 0.588 | 0.576 | 0.591 | 0.584 | 0.594 |
| PGS HR (CI) (p-val) | 1.16 (1.07-1.27) (7.3e-04) | 1.08 (0.98-1.2) (0.10) | 1.22 (1.15-1.30) (1.4e-10) | 1.24 (1.15-1.33) (3.2e-09) | 1.22 (1.11-1.35) (3.8e-05) | 1.27 (1.13-1.42) (7.2e-05) | 1.22 (1.13-1.33) (5.5e-07) | 1.25 (1.13-1.38) (1.3e-05) |
| **Combined Clinical and PGS Model** |  |  |  |  |  |  |  |  |
| C-index | 0.708 | 0.668 | 0.818 | 0.752 | 0.758 | 0.727 | 0.82 | 0.78 |
| CRS HR (CI) (p-val) |  | 2.09 (1.82-2.39) (3.4e-26) |  | 2.78 (2.53-2.05) (6.8e-103) |  | 4.28 (3.32-5.51) (1.7e-29) |  | 4.21 (3.58-4.96) (1.4e-66) |
| PGS HR (CI) (p-val) | 1.23 (1.12-1.34) (6.4e-06) | 1.13 (1.02-1.25) (1.5e-2) | 1.27 (1.19-1.35) (1.8e-14) | 1.27 (1.18-1.36) (5.4e-11) | 1.29 (1.17-1.43) (7.9e-07) | 1.33 (1.18-1.50) (4.5e-06) | 1.29 (1.19-1.39) (4.8e-10) | 1.30 (1.18-1.44) (3.1e-07) |
| **C-index Improvement** | 0.007 | 0.002 | 0.005 | 0.009 | 0.007 | 0.013 | 0.006 | 0.009 |
| **LRT p-value** | 5.85E-06 | 1.43E-02 | 2.01E-14 | 5.75E-11 | 6.65E-07 | 3.84E-06 | 4.29E-10 | 2.93E-07 |

**Supplementary Figure S1:** **Kaplan-meier curves of T2D PGS tertiles by genetic similarity to European ancestry.** Within (A) the total population and (B) a subset of individuals of only EUR ancestry, T2D PGS tertiles separate risk of diabetes onset. However in (C) individuals of non-EUR ancestry, the highest and middle T2D PGS tertiles overlap. Residualizing ancestry bias from the T2D PGS maintains similar separation within (D) the total population and (E) participants of EUR ancestry. (F) Residualization further improves T2D PGS tertile separation in non-EUR participants. Multivariate log-rank test *P* in all analyses < 0.0001.

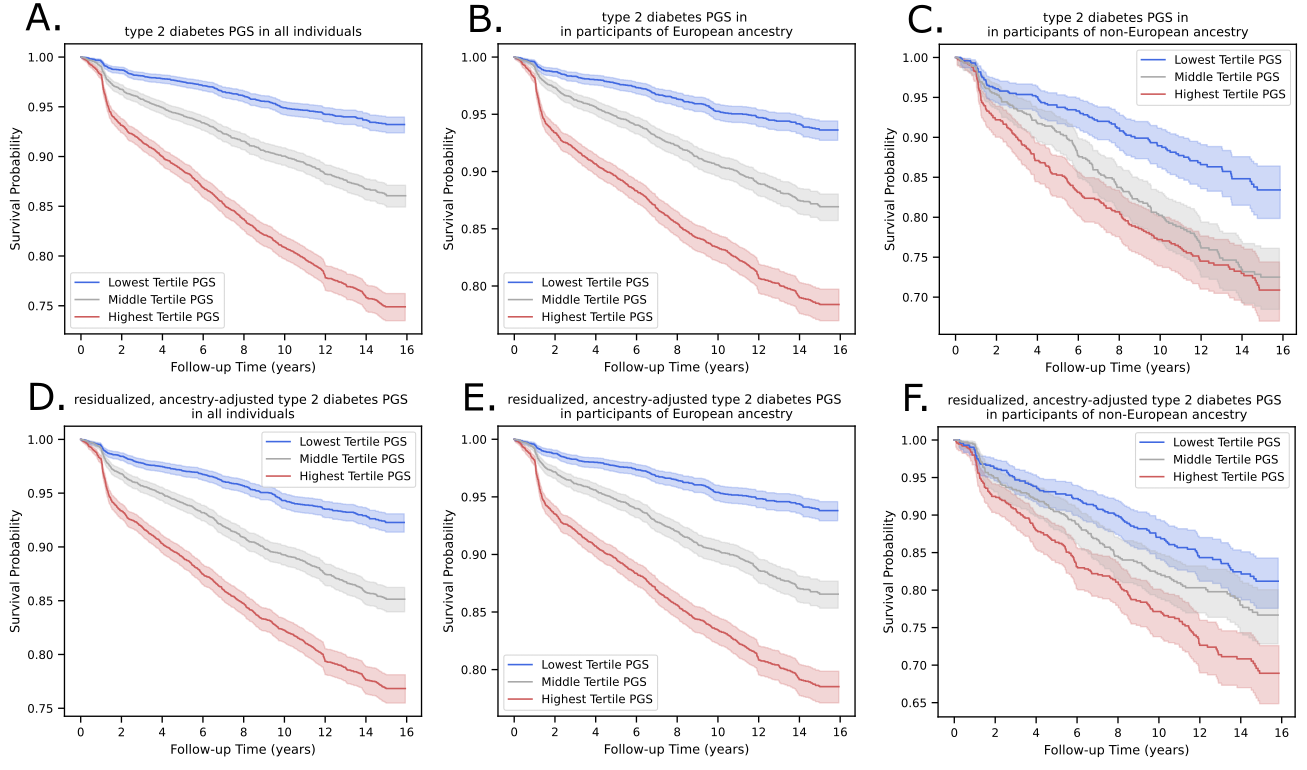
